# The Influence of Precipitation, Runoff, and Temperature on Water Collection Labor in sub-Saharan Africa, 1990-2022

**DOI:** 10.64898/2025.12.22.25342859

**Authors:** Steven Sola

## Abstract

Collecting water in rural sub-Saharan Africa is primarily performed by women. Climate change may affect water resources, damage infrastructure, and limit availability of water for collection. These changes could affect large numbers of rural households and the amount of time they spend collecting water. This study uses national surveys to assess associations between precipitation, surface runoff, and temperature on water collection labor over a 32-year period in rural sub-Saharan Africa. Our outcome was two-way water collection time. Weather data were sourced from ERA5-Land. Information about water source, primary person who collects water, socio-economic status (SES), and Köppen-Geiger climate zone were recorded for each household.

The source population included 1,759,230 households from 183 surveys between 1990 and 2022. The study population included 781,761 rural households that had known water collection times. Among households where the person responsible for collecting water was reported, the greatest number of individuals were adult women (n = 121,777, 70.9%). We utilized multi-level linear regression models with a log-transformed outcome for our analyses. A one-centimeter seven-day cumulative increase in precipitation, and a one-centimeter seven-day cumulative increase in surface runoff, was associated with a 1% reduction in water collection time, while a one-degree Celsius seven-day average increase in temperature was associated with a 2% increase in water collection time.

This study shows that precipitation, surface runoff, and temperature have statistically significant associations with water collection time, although the effect size is small. Further research will need to take a more directed approach to climate change and water collection labor to measure how regional changes to the climate will influence water collection labor. These studies can be used by stakeholders to target infrastructure improvements that would help to alleviate the burden of water collection on rural women and their families as local weather patterns are influenced by climate change.

## Introduction

Access to a clean water source is needed to reduce pathogen exposures that contribute to diarrhea among children under the age of 5 [1,2]. The Joint Monitoring Programme for Water Supply, Sanitation and Hygiene (JMP) is a collaboration between the World Health Organization (WHO) and the United Nations Children’s Fund (UNICEF) that was created in 1990 to report on country, regional, and global estimates of water, sanitation, and hygiene (WASH) indicators. As of 2020, two billion people (26% of the world’s population) lacked safely managed drinking water services, and 2.3 billion people live in water-stressed countries [3]. This issue is particularly striking in sub-Saharan Africa, where only 30% of the population live with a safely managed system, compared to 96% of the population in Europe and North America [3].

Compounding the paucity of safely managed water systems, climate change is expected to cost African nations 2% - 5% of their gross domestic product annually due to climate-related hazards, such as droughts, floods, and cyclones [4]. In fact, some nations allocate up to 9% of their annual budgets to manage climate change [4]. Additionally, climate change can increase the risk of foodborne and waterborne diseases, increase drought, and contribute to more extreme precipitation events leading to flooding and destruction of sanitation infrastructure [5]. The population of sub-Saharan Africa is expected to grow to 1.45 billion people by 2030 from the current estimated population of 1.2 billion people [6]. As access to water may be threatened by the impacts of climate change, the combined effects of this with population growth could amplify individual health issues, such as bodily health, and community issues, such as conflicts over water sources.

When households lack local access to safely managed water systems, travel to collect water for their daily needs is necessary, but this activity also can impact health. Past studies have focused on how water collection labor has impacted women and girls [7–11]. Specifically for sub-Saharan Africa, common issues with water collection labor can include musculoskeletal injuries and fatigue [12–14], school absenteeism [15–17], and negative psychosocial effects [18–20].

Water collection labor has also been shown to be associated with an increase in diarrheal disease [21,22]. It is expected that these negative effects will worsen as climate change continues to shift the precipitation and temperature patterns throughout sub-Saharan Africa, which may impact the burden of water collection labor for households.

As a result, understanding the current and historical status of water collection labor in the context of climate change is critical. While other studies have considered water collection burden within specific countries, few have considered this in the context of climate change and on a broader scale. Improved understanding of water collection labor is important to inform how countries make improvements in their water systems as part of their plans to combat climate change. Therefore, the goal of this work was to expand the knowledge of water collection labor throughout sub-Saharan Africa over the past 30 years in an effort to contribute to the future outlook of water resources and water collection labor.

## Methods

### Study Design

To estimate the association between local weather patterns and water collection labor, we used data from the United States Agency for International Development’s (USAID) Demographic and Health Surveys (DHS) and ERA5-Land weather forecasts from the European Centre for Medium-Range Weather Forecasts (ECMWF) [23].

### Survey Data

DHS surveys are nationally representative household surveys that are funded by USAID and implemented in a partnership between ICF and a host country institution. The first survey was completed in 1985, and USAID has completed over 450 surveys since its inception [24]. Standardized surveys are available; however, host countries typically modify each survey to meet their individual needs. DHS surveys typically have between 5,000 and 30,000 households, with a goal to repeat surveys every five years [25]. Data for each survey are split into separate files depending on the topic. For this analysis, we used the birth recode (BR), household recode (HR), children’s recode (KR), and household member recode (PR) files. Early-year surveys also had a separate file for wealth index (socio-economic status).

We used the term “survey-year” to indicate a particular survey from a particular year (e.g. “South Africa 2016”). Individual files (BR, HR, KR, and PR) from each survey-year were joined together based on key variables linking each file. After the files from each survey-year were joined together, each survey-year was concatenated together to create our dataset. Our final dataset included 1,759,320 households across 183 survey-years. We excluded all households that were considered urban (n = 509,550) or had unknown urbanicity (n = 256,663). According to the DHS surveys, urban areas are typically considered anything bigger than a town, and rural areas are assumed to be countryside [26]. We further excluded households that had missing water collection times (n = 198,977), zero minute water collection times (n = 1,539), or unknown water collection times (n = 45). From the rural households with known collection times, we excluded households that had unrealistic GPS coordinates (n = 10,785), such as households located at a latitude and longitude of (0,0). Our final dataset contained 781,761 households between the years of 1990 and 2022 from 38 different countries across sub-Saharan Africa (see figure 1).

**Figure 1.**
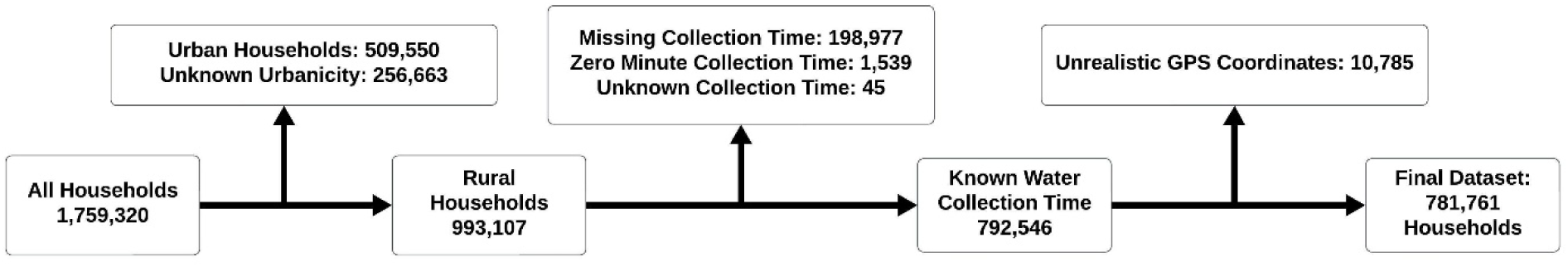
Inclusion of Households in Analysis.

### Weather Data

ERA5-Land is a climate reanalysis dataset at the spatial resolution of 0.1’ x 0.1’ (approximately 9 kms x 9 kms). A total of 11,688 files of daily data were downloaded in NetCDF format from ECMWF. Each daily file included 24 layers, representing each hour of the day, spanning the entire sub-Saharan Africa study region. As a result of ERA5-Land containing data only over land, there were some missing data near coastal and large water body areas. Missing data for each layer in each file were handled by inverse distance weighting based off the four nearest neighbors using Climate Data Operators (CDO) from the Max-Planck-Institut für Meteorologie [27].

Variables of interest for this study included total evaporation (e), surface runoff (sro), sub-surface runoff (ssro), total precipitation (tp), temperature at two meters above ground (t2m), and temperature (skt), which is temperature of the surface of the Earth. ERA5-Land captures these data at 00:00 UTC every day. Survey data in this study were collected from areas that ranged from UTC+0 to UTC+3. Weather data were time zone-shifted to ensure that the weather data were accurately recorded for the specific day of the survey, including lags for previous days before the survey. To accomplish this, we combined the data for the current day and the subsequent day, which gave us data for 48 hours. We then took the time zone and subtracted that number from the beginning, and counted forward 24 hours. The result was a new NetCDF file that accurately incorporated the 24 hours of the survey day.

The variables e, sro, ssro, and tp are considered “accumulation” variables, while t2m and skt are considered “instantaneous” variables. Data for accumulation variables are recorded since the start of the current day, while data for instantaneous variables are recorded for the specific hour. After time zone-shifting each NetCDF, we had the total amount for each of our accumulation variables, and created a 24-hour average for our instantaneous variables.

We utilized the survey date and GPS coordinates for each household present in the final dataset. Dates for the Ethiopian surveys were recorded in the Ethiopian (Ge’ez) calendar. These dates were converted to the Gregorian calendar using the package EthiopianDate [28]. GPS points were matched to the nearest centroid of each grid in the NetCDF files. Köppen-Geiger Zones were recorded for each household based on their GPS coordinates.

### Statistical Analysis

We utilized a mixed-effects linear regression model to assess the influence of weather on water collection time.

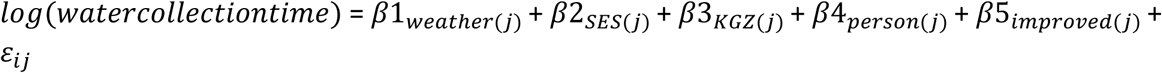

Where:

β_1_ is to the specific weather data of interest

β_2_ is to the SES of the household

β_3_ is to the Koppen-Geiger Zone of the household

β_4_ is to the person who typically collects water for the household

β_5_ is to whether the household has an improved water system

The primary outcome for our analysis was the variable “HV204”, which queries “Time taken to get to the water source for drinking water.” We considered this question as encapsulating both the time to get the water and come back, based on the Guide to DHS Statistics [29] and by observing similar wording in individual country surveys. We log-transformed skewed data when it was the outcome measure to achieve a more normal distribution. When reporting results, we back-transformed the outcome by exponentiating the beta coefficient results from our models.

The primary exposure for our analysis were the seven-day total precipitation accumulation (in centimeters across the entire 0.1’ x 0.1’ [9km x 9km] grid), the seven-day surface runoff accumulation (in centimeters across the entire 0.1’ x 0.1’ [9km x 9km] grid), and the 7-day average temperature (in Celsius). We created three models for this study, with β_1_ representing a specific weather covariate of interest in each model.

Additional covariates include the variable “HV236”, which asks each household who collects the water for the household. Due to differences in coding between survey-years, information for this variable was examined individually for each survey-year and extracted based on each survey-year’s codebook. This allowed us to correct for any changes in coding throughout the years of the survey (see supplemental figure 4).

Information about the primary source of water for each household (HV201) was extracted from each survey. Similar to our primary exposure, the coding for the primary source of water changed depending on the survey-year. Following the same procedure as our primary exposure, we were able to categorize each household’s primary source of water. JMP uses a “ladder” for their drinking water indicators, which separates drinking water availability in five categories.

These categories are: surface water, unimproved (unprotected well or spring), limited (improved source more than 30 minutes from dwelling), basic (improved source less than 30 minutes from dwelling), and safely managed (improved source in/near dwelling and free from contamination) [30]. We used three categories for this study: unimproved (“surface water” and “unimproved” JMP categories), improved (“limited”, “basic”, and “safely managed” JMP categories), and unknown/other (see supplemental figure 5). A Kruskal-Wallis rank sum test was used to assess a statistically significant difference in water collection times among the primary person collecting water for the household, household categories of water source, and households in different Köppen-Geiger Zones.

To assess each household’s SES category, we used the variable HV270. SES with the DHS surveys is constructed with a principal components analysis on a household’s ownership of assets, house construction materials, and access to water and sanitation facilities [31].

Analyses were completed using R 4.2.1 [32]. Replication code is made publicly available on https://github.com/sqsola/ClimateWASH. Data can be obtained through a request to the Demographic and Health Survey team at https://dhsprogram.com/. Permission to download data was received prior to commencing data analyses.

## Results

A total of 1,759,320 households were included in this analysis, encompassing 183 survey-years and 38 countries (see supplemental table 1). The mean water collection time overall was 30.9 minutes. This was further stratified into 993,107 rural households and 509,550 urban households. We excluded 256,663 households due to missing urbanicity information. Rural households, on average, collected water for 33 minutes and urban households, on average, collected water for 23.8 minutes. Households missing urbanicity data collected water for 30.5 minutes on average (see table 1).

**Table 1.** Number and types of households in the study and their average time to collect water, in minutes.

| Type of Household | Households | Average Water Collection Time |
| --- | --- | --- |
|  | N (%) | (Minutes) |
| Rural | 993,107 (56.4) | 33.0 |
| Urban | 509,550 (29) | 23.8 |
| Missing | 256,663 (14.6) | 30.5 |
| Total | 1,759,320 | 30.9 |

### Rural Household Results

We created the following categories for the person who is primarily responsible for collecting water in their household: unknown person (n = 612,370), adult woman (n = 121,777), adult man (n = 21,603), female child under 15 years old (n = 15,790), male child under 15 years old (n = 6,180), any member (n = 2,329), and female and male children combined (n = 1,712). Adult men had the longest average water collection time (37.7 minutes), while female and male children had the shortest average water collection time (19 minutes) (see table 2). The average water collection times among these groups was statistically significantly different ( p < 0.001).

**Table 2.** Primary person responsible for collecting the household water, and their average time to collect water, in minutes.

| Person Responsible for Household Water | Households | Households with Known Water Collector | Average Water Collection Time |
| --- | --- | --- | --- |
|  | N (%) | N (%) | (Minutes) |
| Undetermined Person* | 610,040 (78.0) | - | 33.1 |
| Adult woman | 121,777 (15.6) | 121,777 (70.9) | 32.0 |
| Adult man | 21,603 (2.8) | 21,603 (12.6) | 37.7 |
| Female child under 15 years old | 15,790 (2.0) | 15,790 (9.2) | 29.6 |
| Male child under 15 years old | 6,180 (0.8) | 6,180 (3.6) | 32.5 |
| Any member | 2,329 (0.3) | 2,329 (1.4) | 28.6 |
| Other / Don't Know | 2,330 (0.3) | 2,330 (1.4) | 33.1 |
| Female and male children | 1,712 (0.2) | 1,712 (1.0) | 19.0 |
| Total | 781,761 | 171,721 | 32.9 |

\* Survey question regarding the primary water collector for the household was not included in the core DHS surveys between 2008-2018.

According to the Guide to DHS Statistics, this question was asked as part of the core survey from Phases 1 – 5 (approximately 1985 – 2008). However, this question was dropped in Phases 6 – 7 (approximately 2008-2018), and asked again in Phase 8. As a result, there were a large number of households that did not report the person who was primarily responsible for household water collection. Among those households that had a known person who was primarily responsible for collecting water, adult women collected water in 71.9% of households, followed by adult men, who collected water in 12.8% of households.

A majority of rural households (n = 410,511; 52.51%) had improved water systems. The average water collection time for households with improved systems was 30.6 minutes. Rural households with unimproved systems (n = 368,500; 47.14%) had an average water collection time of 35.5 minutes (see table 3). The average water collection time between these households with different sources of water was statistically significantly different (p < 0.001).

**Table 3.** Categorization of primary water source for the household and the average time to collect water for those households, in minutes.

| Type of Water Source | Households | Average Water Collection Time |
| --- | --- | --- |
|  | N (%) | (Minutes) |
| Improved | 410,511 (52.5) | 30.6 |
| Unimproved | 368,500 (47.1) | 35.5 |
|  | N (%) | (Minutes) |
| Other/Unknown | 2,750 (0.4) | 39.7 |
| Total | 781,761 | 32.9 |

Between 1990 and 2022, there was an increase in the percentage of households who used an improved water source as their primary water source (see figure 2). From 1990-1994, approximately 17.8% of households used an improved source of water, while from 2015-2019, 60.1% of households used an improved source of water (see supplemental table 3). Starting in 2010-2014, there were more households that used an improved water source compared to households that used an unimproved water source.

**Figure 2.**
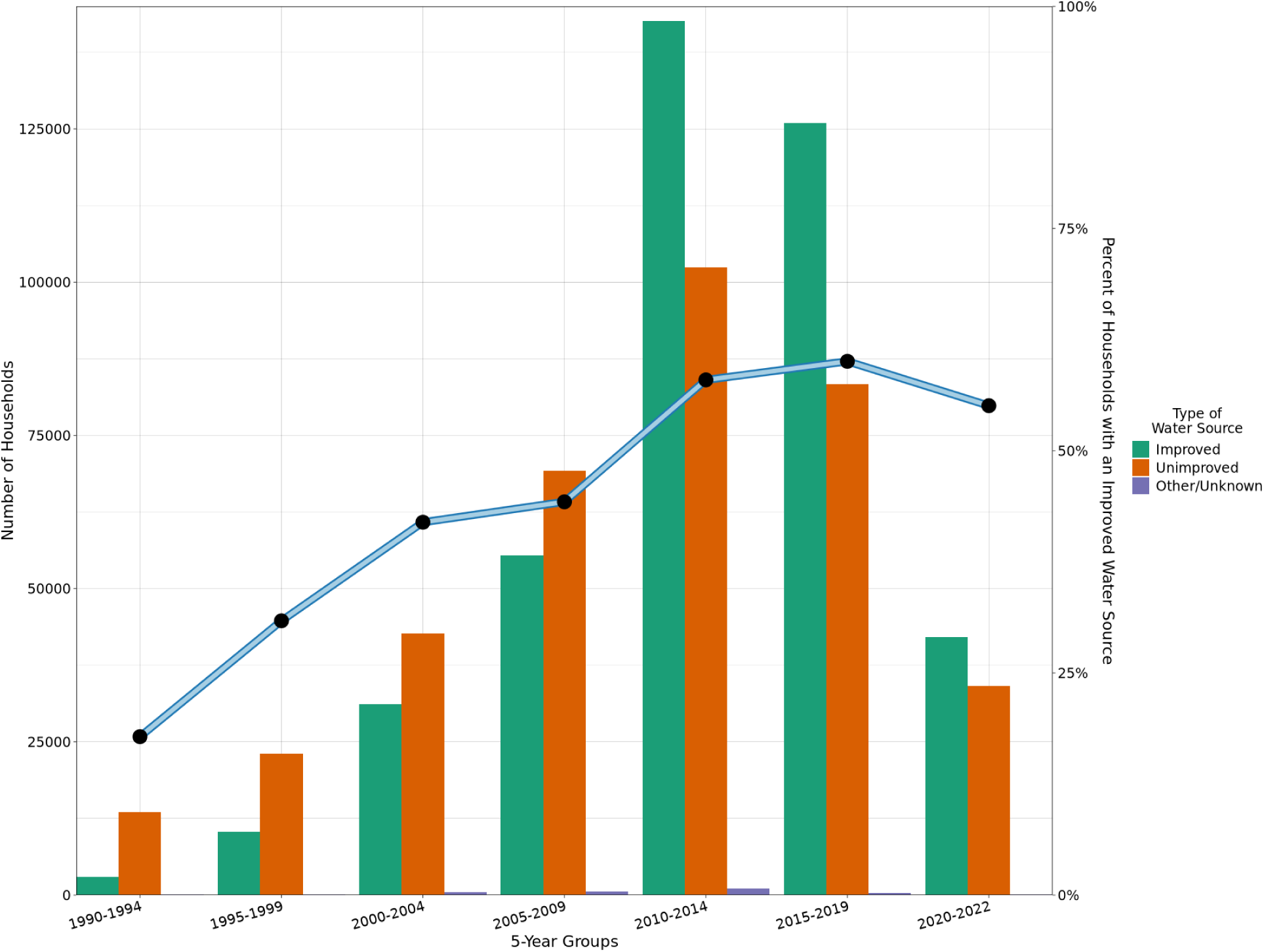
The number of households and their associated water source in five-year groups, along with the percentage of households within the five-year groups with an improved water source.

A majority of the rural households (60.8%) in this study lived in a tropical Köppen-Geiger Zone, where the average water collection time was 32 minutes. A smaller proportion of households (20.3%) lived in temperate areas, with an average water collection time of 31.2 minutes. Most of the other households (18.7%) were located in dry areas, with an average water collection time of 37.9 minutes (see table 4). The average water collection time among these groups was statistically significantly different (p < 0.001).

**Table 4.** Köppen-Geiger climate classification for each household and their average time to collect water, in minutes.

| **Köppen-Geiger Zone** | **Households** | **Average Water Collection Time** |
| --- | --- | --- |
|  | **N (%)** | **(Minutes)** |
| Tropical | 475,560 (60.8) | 32.0 |
| Temperate | 158,961 (20.3) | 31.2 |
| Dry | 146,195 (18.7) | 37.9 |
| Unknown KGZ | 1,045 (0.1) | 20.1 |
| Total | 781,761 | 32.9 |

The results of our three models show a statistically significant association for average water collection time on our three weather covariates, when controlling for SES, Köppen-Geiger Zone, improved water source, and the person responsible for carrying the water (see Table 5). A one-degree Celsius seven-day average increase in Earth’s surface temperature corresponds to a 2% increase in time needed to collect water for the household. Additionally, a one-centimeter seven-day cumulative increase in precipitation, and a one-centimeter seven-day cumulative increase in surface runoff, was associated with a 1% reduction in water collection time, holding all other factors constant.

**Table 5.** Linear mixed-effects model results with average water collection time as the primary outcome and the three primary weather measures as the exposures of interest.

| Covariates | 7-Day Average Temperature (°C) |  | 7-Day Cumulative Surface Runoff (cm) |  | 7-Day Cumulative Precipitation (cm) |  |
| --- | --- | --- | --- | --- | --- | --- |
| | $\beta^1$ | 95% CI<sup>2</sup> | $\beta^1$ | 95% CI<sup>2</sup> | $\beta^1$ | 95% CI<sup>2</sup> |
|  | **1.02** | **1.01, 1.02 \*\*\*** | - | - | - | - |
|  | - | - | **0.99** | **0.98, 1.00 \*** | - | - |
|  | - | - | - | - | **0.99** | **0.99, 1.00 \*\*\*** |
| SES |  |  |  |  |  |  |
| Poorest | Ref | Ref | Ref | Ref | Ref | Ref |
| Poor | **0.95** | **0.94, 0.96 \*\*\*** | **0.95** | **0.94, 0.96 \*\*\*** | **0.95** | **0.94, 0.96 \*\*\*** |
| Middle | **0.94** | **0.92, 0.95 \*\*\*** | **0.93** | **0.92, 0.95 \*\*\*** | **0.93** | **0.92, 0.95 \*\*\*** |
| Rich | **0.91** | **0.90, 0.92 \*\*\*** | **0.91** | **0.89, 0.92 \*\*\*** | **0.91** | **0.89, 0.92 \*\*\*** |
| Richest | **0.87** | **0.85, 0.89 \*\*\*** | **0.86** | **0.85, 0.88 \*\*\*** | **0.86** | **0.85, 0.88 \*\*\*** |
| Köppen-Geiger Zone |  |  |  |  |  |  |
| Dry | Ref | Ref | Ref | Ref | Ref | Ref |
| Temperate | **0.79** | **0.75, 0.84 \*\*\*** | **0.73** | **0.69, 0.77 \*\*\*** | **0.73** | **0.70, 0.77 \*\*\*** |
| Tropical | **0.82** | **0.78, 0.86 \*\*\*** | **0.80** | **0.76, 0.83 \*\*\*** | **0.81** | **0.77, 0.84 \*\*\*** |
| Person Carrying Water |  |  |  |  |  |  |
| Adult man | Ref | Ref | Ref | Ref | Ref | Ref |
| Adult woman | **0.98** | **0.97, 0.99 \*\*** | **0.98** | **0.97, 0.99 \*\*** | **0.98** | **0.97, 0.99 \*\*** |
| Any member | **0.94** | **0.91, 0.98 \*\*\*** | **0.94** | **0.91, 0.98 \*\*\*** | **0.94** | **0.91, 0.98 \*\*** |
| Female and male children | 1.01 | 0.97, 1.05 | 1.01 | 0.97, 1.05 | 1.01 | 0.97, 1.05 |
| Female child under 15 years old | 1.00 | 0.98, 1.01 | 1.00 | 0.98, 1.01 | 1.00 | 0.98, 1.01 |
| Male child under 15 years old | 1.02 | 1.00, 1.04 | 1.02 | 1.00, 1.04 | 1.02 | 1.00, 1.04 |
| Improved Water Source |  |  |  |  |  |  |
| Improved | Ref | Ref | Ref | Ref | Ref | Ref |
| Other/Unknown | **0.88** | **0.83, 0.94 \*\*\*** | **0.88** | **0.83, 0.94 \*\*\*** | **0.88** | **0.83, 0.94 \*\*\*** |
| Unimproved | **1.16** | **1.14, 1.17 \*\*\*** | **1.16** | **1.14, 1.17 \*\*\*** | **1.16** | **1.14, 1.17 \*\*\*** |

<sup>1</sup> Exponentiated beta coefficient from a log-transformed dependent variable. Interpret on a multiplicative scale, not on an additive scale, such that a result of less than 1 indicates a decrease in relative water collection time and a result of more than 1 indicates an increase in relative water collection time.
<sup>2</sup> CI = Confidence Interval
\* p<0.05, \*\* p<0.01, \*\*\* p<0.001

There was a decrease in the amount of time taken to collect water as a household’s SES group increases, with the richest households reported taking approximately 13% less time to collect water compared to the poorest households. Households living in temperate climate zones reported taking 21% - 27% less time to collect water, and households living in tropical climate zones reported taking 18%-20% less time to collect water, compared to households living in dry climate zones.

Households in which women were the primary household water collector reported 2% less time needed to collect water, and households where any member could be the primary household water collector reported 6% less time needed to collect water, compared to households that reported men as the primary household water collector. Households where children were the primary household water collector had statistically similar water collection times compared to households where men were the primary household water collector. Households that reported collecting water from unimproved water sources had a 16% increase in the time needed to collect water compared to households that reported collecting water from improved water sources.

## Discussion

This research found that increases in precipitation and surface runoff were associated with decreases in water collection time, after considering a household’s SES, climate zone, who collects the water, and whether the water source is improved or unimproved. Given that the evaluation considered water collection labor over the past 32 years in sub-Saharan Africa, these results can be used to inform understanding of the burden of water collection labor as climate change progresses. Although most households did not report who collected the water, when they did report, women were the primary source of water collection labor, which is concordant with prior studies [7,9,30]. As such, women may be most likely to be affected by climate change in the context of water collection labor, given the potential of worsening water scarcity throughout the subcontinent.

These findings replicate and expand prior research efforts to understand who collects water and how water collection may be impacted by weather and climate. In 2016, a study by Graham et al. used data from 24 countries in the DHS Program and UNICEF’s Multiple Indicators Cluster Surveys (MICS) between the years 2005-2012 to assess water collection labor [7]. The authors found that adult women were predominantly responsible for the water collection, and in 10 countries, adult women were the primary water collector in more than 75% of the households surveyed. Similar results were seen in our study, where we found that women were responsible for collecting water in 71.9% of households where the person responsible for collecting water was identified. Another similarity was that female children were more likely to be water collectors than male children.

Other studies have examined how precipitation may be associated with the time needed to collect water. A preprint from Paulos et al. in 2023 used data from 104 surveys and 34 countries from the DHS program to study the association between temperature and precipitation on water collection times in sub-Saharan Africa [33]. They found that a 1 cm increase in precipitation is associated with a decrease in walk time of between 0.3 minutes to 5.1 minutes, depending on the time period. They also found that a one-degree Celsius increase in temperature is associated with an increased collection time of between 0.43 minutes to 1.21 minutes, depending on the time period. These finding are concordant with our results, where we also found that an increase in precipitation led to a decrease in water collection time, and an increase in temperature led to a small increase in water collection time.

This study builds upon the previous studies by using a larger dataset and a single source of climate data at a fine spatial resolution. The current study included 183 survey-years from 1990-2022, and included countries that previous studies have excluded, such as Ethiopia. (DHS data from Ethiopia uses the Ethiopian (Ge’ez) calendar, which is approximately seven to eight years behind the Gregorian calendar that much of the rest of the world uses.) This study also incorporates the concept of surface runoff into statistical models, something that previous studies have overlooked. Surface runoff is the measurement of the amount of water that is accumulated on the surface of a particular area, and this measurement can be used as an indicator of drought or flood [34]. Areas with a high amount of surface runoff could indicate areas where water collection is easier, especially for households that still rely on surface water for their daily needs. Alternatively, areas with a low amount of surface runoff could indicate areas where water collection is more difficult. The use of surface runoff could help us to more fully understand the influences of local weather patterns on water collection labor. This association may also be modified by whether a household collects from an unimproved source (such as surface water) compared to an improved source (such as a borehole, which is less influenced by surface runoff).

A strength of this study was the use of ERA-5 Land climate data, which was available for dates since the start of the DHS program, and has a finer spatial resolution (0.1’ x 0.1’) than other common sources of weather data, such as Climate Hazards Group InfraRed Precipitation with Station data (CHIRPS) at a spatial resolution of only 0.25’ x 0.25’. ERA-5 Land includes not only precipitation data, but also surface runoff and Earth’s temperature using the same source dataset. This helped to reduce any potential errors from needing to use multiple datasets for weather data, which would have introduced errors due to differing methods of measurement and differing spatial resolutions between datasets.

There were some limitations to this analysis. There were a large number of unknown people who collected water for their household (78.3%) due to the DHS program not collecting that information on their standard questionnaire for approximately 10 years. Additionally, the questionnaire does not make clear whether the outcome of interest is the two-way water collection time, or only one-way water collection time. While the questionnaire indicates this should be two-way collection time, other analyses have interpreted it as a one-way collection time [31, 32]. SES in this analysis also should be interpreted with caution. In the DHS data, SES is constructed with regard to all households in the survey-year. As a result, most rural households are at the lower end of the SES measure, and most urban households are at the higher end of the SES measure. DHS recently has started stratifying SES measures based on urbanicity, but using this measure would have further restricted our analysis. Further caution is warranted due to the long time frame of this study (1990-2022), as well as the large number of surveys that were included in this analysis. Each survey that has been implemented includes both sampling and nonsampling errors. Sampling errors can be ameliorated by statistics to account for the sample of people selected for the survey and their representativeness of the source population. Nonsampling errors, such as data collection, data processing, and question misunderstanding, cannot be easily accounted for by statistics. Each survey used in this study has a report in the DHS website that details these errors, but these corrections were not implemented in this study. Finally, the interpretability of the weather variables used in this analysis may be difficult, as both the total precipitation and surface runoff measurements are averaged over their representative 0.1’ x 0.1’ grid locations, and aren’t indicative of local measurements in their locations. A household within a specific grid could be exposed to different weather patterns compared to other households within the same grid.

This research uses a dataset that is rich in both demographic and climate data. Future studies can use this dataset to evaluate individual countries, general areas of sub-Saharan Africa, or specific time frames to answer questions not only related to water collection, but also other topics, such as maternal and child health, poverty, and sanitation. An example would be a study by Dimitrova et al. in 2022 used DHS data and found that anomalous wet and dry conditions could increase the risk of cough, fever and diarrhea, depending on the climate zone [35]. This work can be supplemented with qualitative studies that assess how climate change may impact water collection practices in the future, similar to the work undertaken by Bose et. al in their paper looking at seasonality and water security in rural Gambia [36]. This research can help to explain the importance for including climate change into discussions of water collection labor, especially because climate change will influence the amount of freshwater that is available reliably to collect. This research provides evidence to support greater attention on how the impending climate change crisis will adversely impact women and girls, and the benefits that an increase in spending on water infrastructure can be implemented in ways that to promote gender equity throughout the sub-continent.

## Data Availability

All data produced in the present study are available upon reasonable request to the authors

## Acknowledgements

This work was carried out at the Advanced Research Computing at Hopkins (ARCH) core facility (rockfish.jhu.edu), which is supported by the National Science Foundation (NSF) grant number OAC1920103.

---

**Supplement Figure 1.**
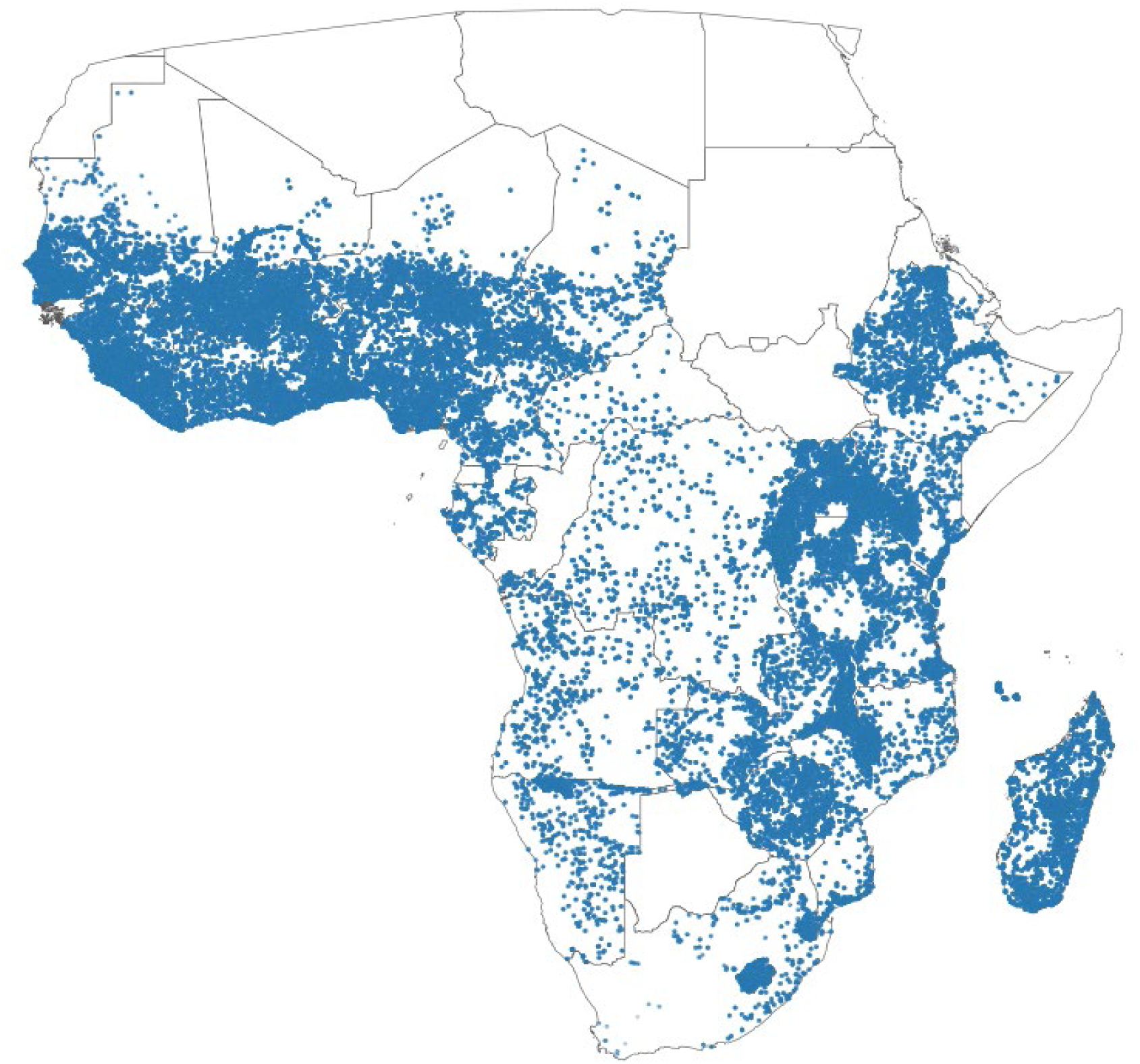
Locations of rural households included in this analysis.

**Supplemental Figure 2.**
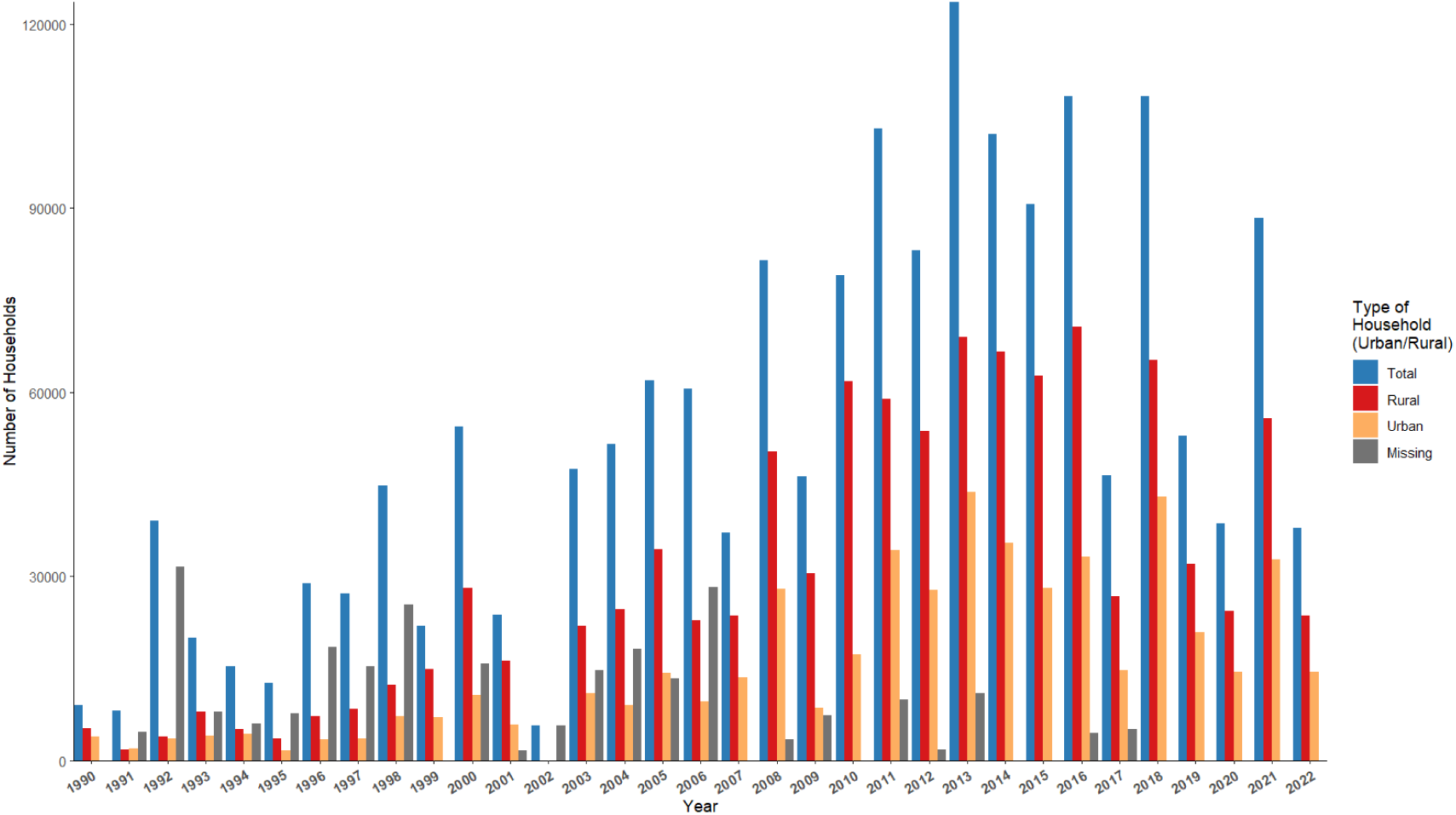
Total number of households by year and their associated urbanicity.

**Supplemental Figure 3.**
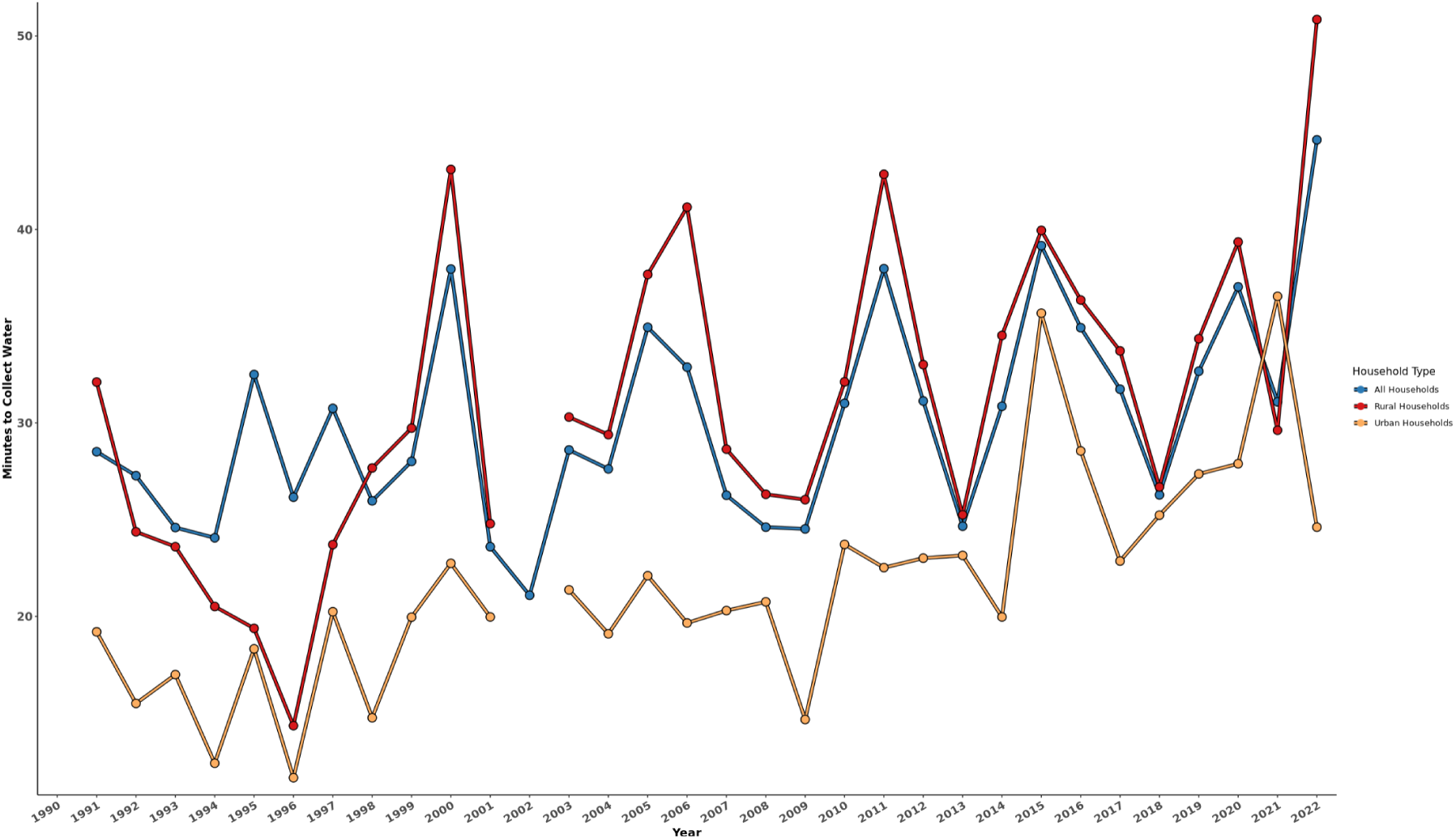
Average water collection time by year and their related urbanicity.

**Supplemental Figure 4.**
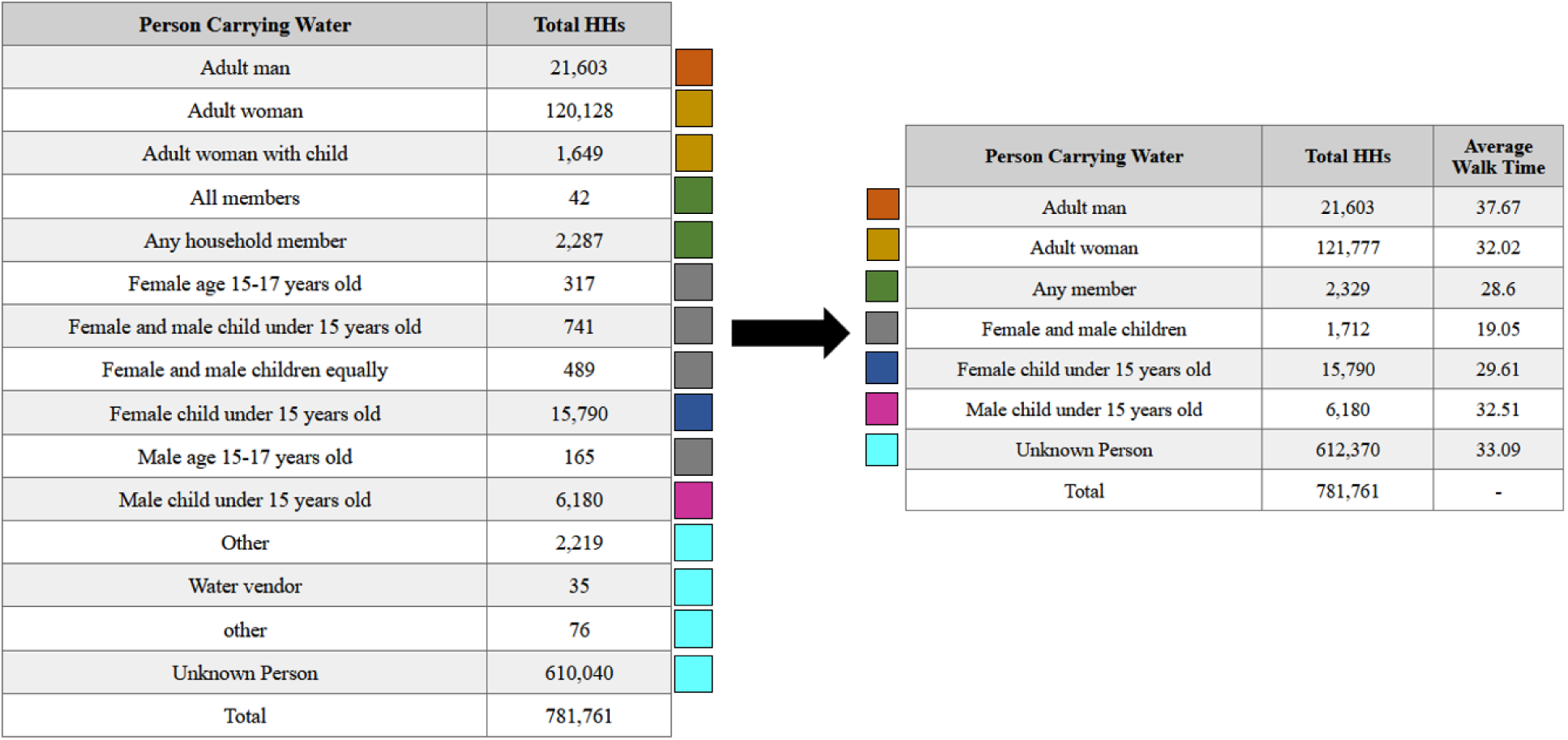
Categorization of the covariate “person carrying water”.

**Supplemental Figure 5.**
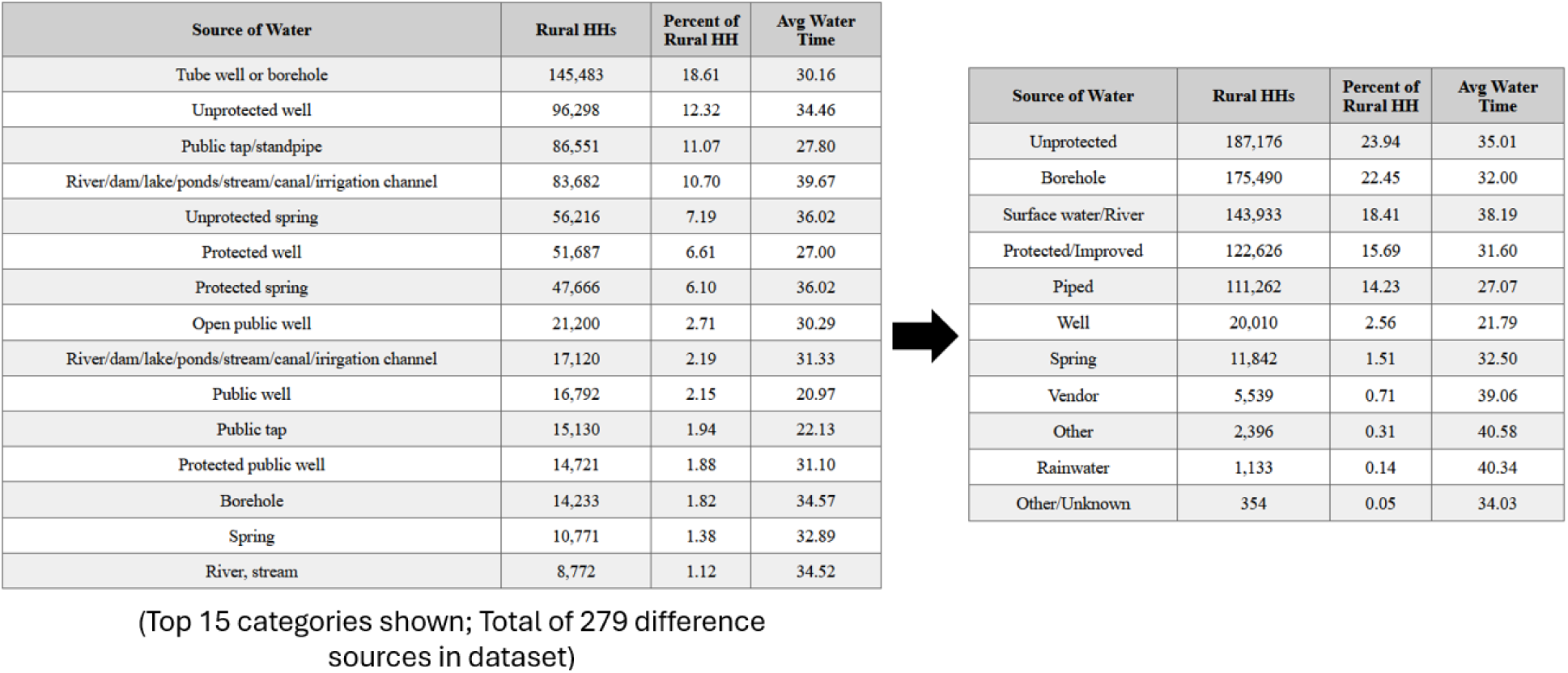
Categorization of sources of water, the percentage of rural households for each source, and the average water collection time for each source.

**Supplement Table 1.** Countries and survey-years used in this analysis.

| Country | Year |
| --- | --- |
| Angola | 2006-2007, 2011, 2015-2016 |
| Benin | 1996, 2001, 2006, 2011-2012, 2017-2018 |
| Burkina Faso | 1993, 1998-1999, 2003, 2010, 2014, 2017-2018, 2021 |
| Burundi | 2010, 2012, 2016-2017 |
| Cameroon | 1991, 1998, 2004, 2011, 2018 |
| Central African Republic | 1994-1995 |
| Chad | 1996-1997, 2004, 2014-2015 |
| Comoros | 1996, 2012 |
| Congo | 2005, 2009, 2011-2012 |
| Democratic Republic of the Congo | 2007, 2013-2014 |
| Cote d'Ivoire | 1994, 1998-1999, 2005, 2011-2012, 2021 |
| Eswatini | 2006-2007 |
| Ethiopia | 2000, 2005, 2011, 2016, 2019 |
| Gabon | 2000, 2012, 2019-2021 |
| Gambia | 2013, 2019-2020 |
| Ghana | 1993, 1998, 2003, 2008, 2014, 2016, 2019 |
| Guinea | 1999, 2005, 2012, 2018 |
| Kenya | 1993, 1998, 2003, 2008-2009, 2014, 2015, 2020, 2022 |
| Lesotho | 2004, 2009, 2014 |
| Liberia | 2007, 2009, 2011, 2013, 2016, 2019-2020 |
| Madagascar | 1992, 1997, 2003-2004, 2008-2009, 2011, 2013, 2016, 2021 |
| Malawi | 1992, 2000, 2004, 2010, 2012, 2014, 2015-2016, 2017 |
| Mali | 1995-1996, 2001, 2006, 2012-2013, 2015, 2018, 2021 |
| Mauritania | 2019-2021 |
| Mozambique | 1997, 2003, 2011, 2018 |
| Namibia | 1992, 2000, 2006-2007, 2013 |
| Niger | 1992, 1998, 2006, 2012, 2021 |
| Nigeria | 1990, 2003, 2008, 2010, 2013, 2015, 2018, 2021 |
| Rwanda | 1992, 2000, 2005, 2007-2008, 2010, 2013, 2014-2015, 2017, 2019-2020 |
| Sao Tome and Principe | 2008-2009 |
| Senegal | 1992-1993, 1997, 2005, 2006, 2008-2009, 2010-2011, 2012-2014, 2015-2016, 2017, 2018, 2019, 2020-2021 |
| Sierra Leone | 2008, 2013, 2016, 2019 |
| South Africa | 1998, 2016 |
| Tanzania | 1991-1992, 1996, 1999, 2003-2004, 2004-2005, 2007-2008, 2010, 2011-2012, 2015-2016, 2017 |
| Togo | 1998, 2013-2014, 2017 |
| Uganda | 1995, 2000-2001, 2006, 2009, 2011, 2014-2015, 2016, 2018-2019 |
| Zambia | 1992, 1996, 2001-2002, 2007, 2014-2014, 2018 |
| Zimbabwe | 1994, 1999, 2005-2006, 2010-2011, 2015 |

**Supplement Table 2.** Total number of households and their average water collection times, stratified on urbanicity.

| Year | Total HHs | Mean Collection Time | Rural HHs | Rural Mean Collection Time | Urban HHs | Urban Mean Collection Time | HH Missing Urbanicity | Missing Mean Collection Time |
| --- | --- | --- | --- | --- | --- | --- | --- | --- |
| 1990 | 8,999 |  | 5,130 |  | 3,869 |  | 0 |  |
| 1991 | 8,124 | 28.51 | 1,715 | 32.11 | 1,823 | 19.20 | 4,586 | 29.58 |
| 1992 | 39,070 | 27.27 | 3,906 | 24.37 | 3,594 | 15.50 | 31,570 | 28.47 |
| 1993 | 19,960 | 24.59 | 7,980 | 23.60 | 4,030 | 16.99 | 7,950 | 28.47 |
| 1994 | 15,341 | 24.06 | 5,106 | 20.51 | 4,251 | 12.41 | 5,984 | 33.25 |
| 1995 | 12,615 | 32.50 | 3,533 | 19.38 | 1,532 | 18.32 | 7,550 | 40.46 |
| 1996 | 28,877 | 26.16 | 7,175 | 14.35 | 3,329 | 11.66 | 18,373 | 30.23 |
| 1997 | 27,199 | 30.75 | 8,335 | 23.71 | 3,608 | 20.24 | 15,256 | 36.53 |
| 1998 | 44,824 | 25.97 | 12,322 | 27.67 | 7,178 | 14.76 | 25,324 | 26.89 |
| 1999 | 21,956 | 28.01 | 14,903 | 29.73 | 7,053 | 19.96 | 0 |  |
| 2000 | 54,357 | 37.95 | 28,007 | 43.10 | 10,558 | 22.74 | 15,792 | 34.03 |
| 2001 | 23,711 | 23.60 | 16,256 | 24.79 | 5,841 | 19.97 | 1,614 | 21.04 |
| 2002 | 5,619 | 21.09 | 0 |  | 0 |  | 5,619 | 21.09 |
| 2003 | 47,500 | 28.60 | 21,845 | 30.30 | 10,931 | 21.37 | 14,724 | 29.56 |
| 2004 | 51,573 | 27.62 | 24,546 | 29.39 | 8,955 | 19.10 | 18,072 | 28.27 |
| 2005 | 61,905 | 34.94 | 34,448 | 37.67 | 14,167 | 22.10 | 13,290 | 33.74 |
| 2006 | 60,604 | 32.88 | 22,827 | 41.15 | 9,543 | 19.66 | 28,234 | 28.14 |
| 2007 | 37,031 | 26.26 | 23,548 | 28.64 | 13,483 | 20.30 | 0 |  |
| 2008 | 81,542 | 24.61 | 50,336 | 26.31 | 27,876 | 20.75 | 3,330 | 22.34 |
| 2009 | 46,221 | 24.52 | 30,472 | 26.03 | 8,447 | 14.67 | 7,302 | 24.28 |
| 2010 | 79,078 | 31.01 | 61,832 | 32.12 | 17,246 | 23.72 | 0 |  |
| 2011 | 103,016 | 37.97 | 58,953 | 42.85 | 34,164 | 22.52 | 9,899 | 40.71 |
| 2012 | 83,132 | 31.13 | 53,618 | 33.01 | 27,781 | 23.01 | 1,733 | 42.07 |
| 2013 | 123,650 | 24.66 | 68,953 | 25.25 | 43,714 | 23.15 | 10,983 | 26.16 |
| 2014 | 102,039 | 30.86 | 66,637 | 34.52 | 35,402 | 19.97 | 0 |  |
| 2015 | 90,620 | 39.16 | 62,606 | 39.95 | 28,014 | 35.67 | 0 |  |
| 2016 | 108,266 | 34.92 | 70,627 | 36.35 | 33,202 | 28.55 | 4,437 | 29.10 |
| 2017 | 46,469 | 31.74 | 26,770 | 33.72 | 14,658 | 22.86 | 5,041 | 32.98 |
| 2018 | 108,224 | 26.28 | 65,234 | 26.69 | 42,990 | 25.23 | 0 |  |
| 2019 | 52,839 | 32.67 | 31,928 | 34.35 | 20,911 | 27.36 | 0 |  |
| 2020 | 38,660 | 37.03 | 24,264 | 39.35 | 14,396 | 27.89 | 0 |  |
| 2021 | 88,388 | 31.09 | 55,713 | 29.62 | 32,675 | 36.54 | 0 |  |
| 2022 | 37,911 | 44.63 | 23,582 | 50.85 | 14,329 | 24.61 | 0 |  |

**Supplement Table 3.** Number of households and their primary source of water by 5-year groups.

| **Water Source** | **5-Year Categories** | **Number of Households** | **Percent per 5-Year Group** |
| --- | --- | --- | --- |
| Improved | 1990-1994 | 2,962 | 0.1783 |
| Unimproved | 1990-1994 | 13,600 | 0.8184 |
| Other/Unknown | 1990-1994 | 55 | 0.0033 |
| Improved | 1995-1999 | 10,357 | 0.3086 |
| Unimproved | 1995-1999 | 23,101 | 0.6884 |
| Other/Unknown | 1995-1999 | 101 | 0.0030 |
| Improved | 2000-2004 | 31,154 | 0.4196 |
| Unimproved | 2000-2004 | 42,682 | 0.5748 |
| Other/Unknown | 2000-2004 | 419 | 0.0056 |
| Improved | 2005-2009 | 55,407 | 0.4425 |
| Unimproved | 2005-2009 | 69,215 | 0.5528 |
| Other/Unknown | 2005-2009 | 588 | 0.0047 |
| Improved | 2010-2014 | 142,636 | 0.5797 |
| Unimproved | 2010-2014 | 102,352 | 0.4160 |
| Other/Unknown | 2010-2014 | 1,060 | 0.0043 |
| Improved | 2015-2019 | 125,929 | 0.6006 |
| Unimproved | 2015-2019 | 83,365 | 0.3976 |
| Other/Unknown | 2015-2019 | 387 | 0.0018 |
| Improved | 2020-2022 | 42,066 | 0.5507 |
| Unimproved | 2020-2022 | 34,185 | 0.4475 |
| Other/Unknown | 2020-2022 | 140 | 0.0018 |

